# Cardiac age prediction using graph neural networks

**DOI:** 10.1101/2023.04.19.23287590

**Authors:** Marco H. de A. Inácio, Mit Shah, Mina Jafari, Nairouz Shehata, Qingjie Meng, Wenjia Bai, Axel Gandy, Ben Glocker, Declan P. O’Regan

## Abstract

The function of the human heart is characterised by complex patterns of motion that change throughout our lifespan due to accumulated damage across biological scales. Understanding the drivers of cardiac ageing is key to developing strategies for attenuating age-related processes. The motion of the surface of the heart can be conceived as a graph of connected points in space moving through time. Here we develop a generalisable framework for modelling three-dimensional motion as a graph and apply it to a task of predicting biological age. Using sequences of segmented cardiac imaging from 5064 participants in UK Biobank we train a graph neural network (GNN) to learn motion traits that predict healthy ageing. The GNN outperformed (mean absolute error, MAE = 4.74 years) a comparator dense neural network and boosting methods (MAE = 4.90 years and 5.08 years, respectively). We produce human-intelligible explanations of the predictions and using the trained model we also assess the effect of hypertension on biological age. This work shows how graph representations of complex motion can efficiently predict biologically meaningful outcomes.

## Introduction

Graphs enable representation of complex data and their interactions in a perceptually meaningful way.^1^ Their relevance to understanding data sampled from biomedical domains offers new approaches for learning and inference in complex real-world systems. Such approaches have been applied to rich sources of heterogeneous high-dimensional clinical data to model physical or functional connectivity in non-Euclidean representations of body systems. For instance, *in vivo* imaging inherently manifests a graph structure comprising pixels that can be represented as nodes and the connections between them as edges with several applications emerging across neuroscience domains.^2^ In contrast to brain imaging, cardiovascular imaging also encodes a temporal dimension which can be represented by dynamic coordinate data derived from shape modelling.^3,4^ Recently, graph representations of synthetic cardiac motion have enabled emulation of such multi-scale systems where the dynamic topology of nodes embeds biomechanically-relevant features.^5^ Here we propose a generalisable framework for modelling three-dimensional (3D) motion of connected points in space as a graph and apply it to a predictive task in human health using information abstracted from cardiac imaging.

Cardiovascular disease is a leading cause of death globally, and ageing is a primary risk factor for its development and progression. As the heart ages it undergoes changes in volume, mass and function as a consequence of increased mechanical stiffness, impaired contractility, and altered tissue energetics. Global parameters derived from cardiovascular imaging vary with age and can be used to predict the difference between chronological age and biological age by reference to a normative population.^6^ This approach allows the identification of factors which may accelerate or attenuate age-related processes. However, such models using hand-crafted features have relatively weak predictive power as the full spectrum of multi-scale spatio-temporal motion traits associated with cardiac ageing are unknown. From a computational perspective this problem is one of learning the relationship between the motion of connected points in space and a continuous outcome, which may be tractable through supervised learning of a graph model derived from time-resolved imaging.

Our input data were cardiac magnetic resonance (CMR) imaging studies in UK Biobank (UKB), providing consistent acquisitions at scale. We performed segmentation and motion tracking to build a dynamic three-dimensional mesh encoding variation in shape and function of the heart with respect to age. These time-resolved meshes were transformed into graph structures and passed to a graph neural network (GNN) which was trained on a cohort of healthy subjects to predict age. We assessed predictive performance in comparison to other methods, and also evaluated the effect of hypertension on biological ageing. We show that graph representations of three-dimensional motion data provide efficient predictions of human health with intelligible explanations.

## Results

### Study Overview

The UKB is a prospective longitudinal study that has recruited approximately 500,000 participants from the United Kingdom with deep genetic and phenotypic data.^7^ A sub-study of UKB invited participants for CMR to assess cardiac volumes and function acquired using a standard protocol.^8^ We first partitioned this dataset by selecting 5064 “healthy” participants and 1330 participants diagnosed with hypertension. Baseline characteristics of both groups are shown in Table 1. We then performed image segmentation and motion tracking to represent the heart as a moving three dimensional mesh. We next built a graph representation of cardiac motion for training a GNN to predict age. The trained model was subsequently used to predict age in the 1330 participants with hypertension. Using the age prediction, we then calculate the “age-delta” in participants with hypertension, quantifying the difference between true chronological age and predicted biological age. An outline of the main steps for the analysis are shown in Figure 1.

**Table 1.** Participant characteristics. A summary of baseline demographic, clinical and imaging characteristics in the UK Biobank healthy and hypertensive groups used in our study, stratified by sex. Abbreviations: LV, left ventricle; CMR, Cardiac magnetic resonance.

| Characteristic | Healthy participants |  | Hypertension participants |  |
| --- | --- | --- | --- | --- |
|  | Females, N = 3,149<sup>1</sup> | Males, N = 1,915<sup>1</sup> | Females, N = 718<sup>1</sup> | Males, N = 612<sup>1</sup> |
| **Baseline characteristics** |  |  |  |  |
| Age at CMR (years) | 61 (55, 67) | 62 (56, 68) | 65 (59, 70) | 65 (59, 70) |
| Race, Caucasian, n (%) | 2,824 (90%) | 1,712 (89%) | 628 (87%) | 534 (87%) |
| Body mass index (kg/m<sup>2</sup>) | 23.67 (21.95, 25.67) | 24.92 (23.20, 26.73) | 24.99 (23.05, 27.18) | 26.17 (24.49, 27.80) |
| Systolic blood pressure (mmHg) | 122 (102, 136) | 130 (110, 142) | 138 (116, 154) | 142 (111, 155) |
| Diastolic blood pressure (mmHg) | 71 (57, 78) | 74 (62, 82) | 78 (66, 86) | 82 (63, 88) |
| Pulse rate (bpm) | 70 (63, 78) | 65 (58, 73) | 72 (65, 81) | 67 (61, 77) |
| **CMR parameters** |  |  |  |  |
| LV end-diastolic volume (ml) | 127 (113, 141) | 167 (149, 188) | 126 (114, 141) | 164 (146, 185) |
| LV end-systolic volume (ml) | 49 (42, 57) | 70 (60, 81) | 48 (40, 56) | 67 (57, 80) |
| LV stroke volume (ml) | 77 (68, 87) | 97 (86, 110) | 78 (69, 88) | 96 (85, 109) |
| LV ejection fraction (%) | 61.2 (57.5, 64.8) | 58.2 (54.6, 61.7) | 61.9 (57.8, 65.9) | 58.8 (54.9, 62.7) |
| LV cardiac output (L/min) | 4.73 (4.16, 5.44) | 5.62 (4.97, 6.43) | 4.94 (4.36, 5.70) | 5.95 (5.30, 6.80) |
| LV mass (g) | 67 (61, 74) | 97 (87, 109) | 72 (65, 80) | 101 (90, 113) |
| Peak longitudinal strain rate (s<sup>-1</sup>) | 1.80 (1.42, 2.21) | 1.63 (1.28, 1.99) | 1.55 (1.23, 1.97) | 1.41 (1.12, 1.79) |
| Peak radial strain rate (s<sup>-1</sup>) | -6.37 (-7.53, -5.16) | -5.72 (-6.98, -4.57) | -6.01 (-7.31, -4.39) | -5.32 (-6.49, -4.26) |

<sup>1</sup>Median (IQR); n (%)

**Figure 1.**
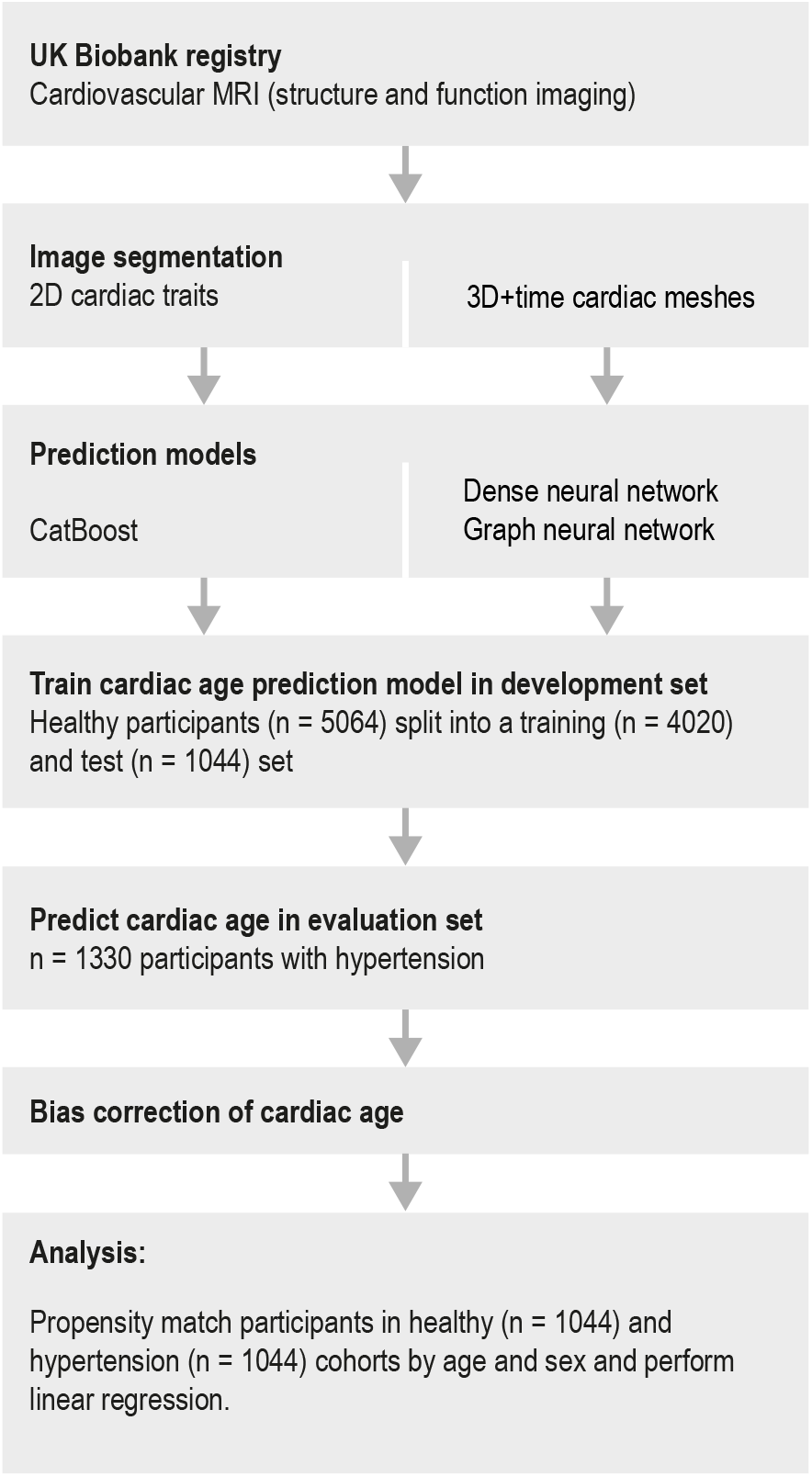
Study flowchart. A summary of the main steps in our analysis of cardiac motion meshes to prediction of age-delta and analysis. MRI, Magnetic Resonance Imaging

### Image phenotyping

We assessed left ventricular volumes and mass, as well as measures of systolic and diastolic function, using segmentation of 319,650 individual cine images by a fully convolutional network (FCN) (Figures 2 and 3A). In total 104 conventional image-derived phenotypes were collected for each participant (selected parameters in Table 1 and Figure 4), which were subsequently used as tabular data for age prediction in a comparative model using gradient boosting. Compared to healthy controls the hypertensive group had higher body mass index (25.5 kg/m^2^ vs 24.3 kg/m^2^, *P* = 2.7 × 10^-49^), cardiac output (5.6 L/min vs 5.2 L/min, *P* = 8.5 × 10^-19^), and left ventricular mass (86.8 g vs 79.5 g, *P* = 1.1 × 10^-28^), but decreased peak strain rate measurements (peak longitudinal strain rate: 1.5s^-1^ vs 1.8s^-1^, *P* = 2.9 × 10^-36^; and peak radial strain rate: -5.5s^-1^ vs -6.1s^-1^, *P* = 1.3 × 10^-14^) (Supplementary Table 1). As inputs to a second comparative deep learning model, we derived time-resolved 3D motion models of the heart using non-rigid image registration between cine images. Aligning data to an atlas template allowed motion traits to be aggregated in a standard coordinate space, which demonstrated complex spatio-temporal differences in motion fields between health states (Figure 3B).

**Figure 2.**
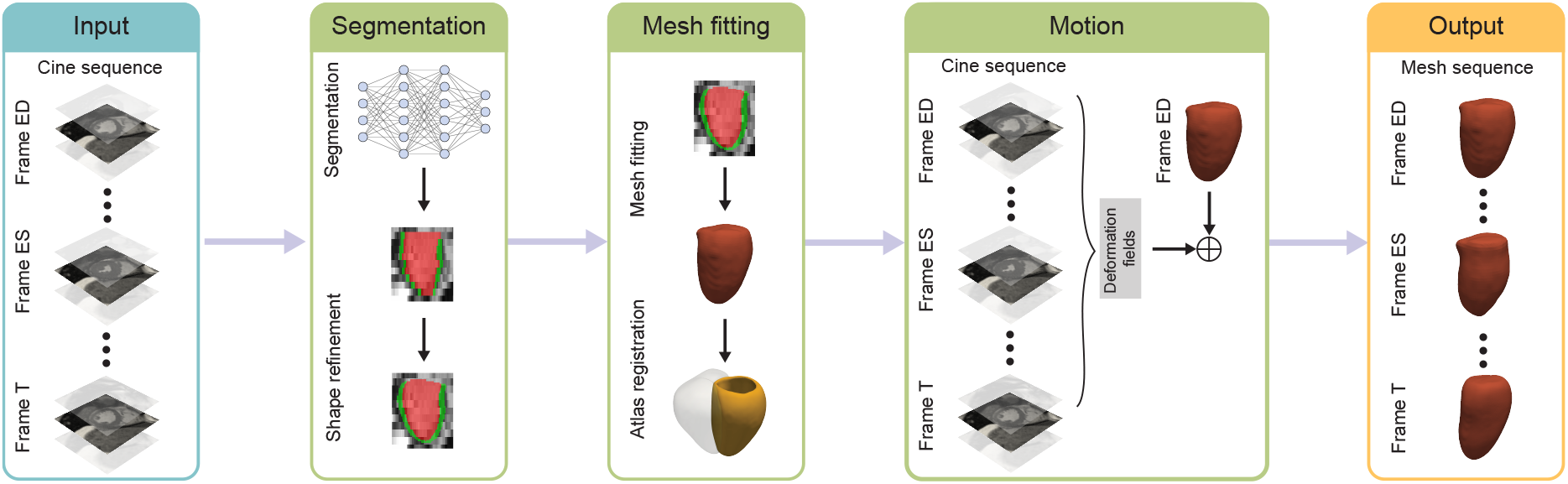
Overview of image processing framework. Short axis cine sequences of the heart from cardiac magnetic resonance imaging (3 sections from the *≈*10 acquired slices shown) are segmented and shape refined using a multitask network (red - blood pool, green - myocardium). Mesh fitting of the left ventricle is performed using marching cubes and an atlas template registered to establish anatomical correspondence. A deformation field is obtained from the cine images and motion encoded in the mesh models. The output are left ventricular meshes in 5064 participants with 50 temporal phases where coordinates of each vertex are in a standard reference space. (ED, end diastole; ES, end systole; T, temporal phase 1 - 50.)

**Figure 3.**
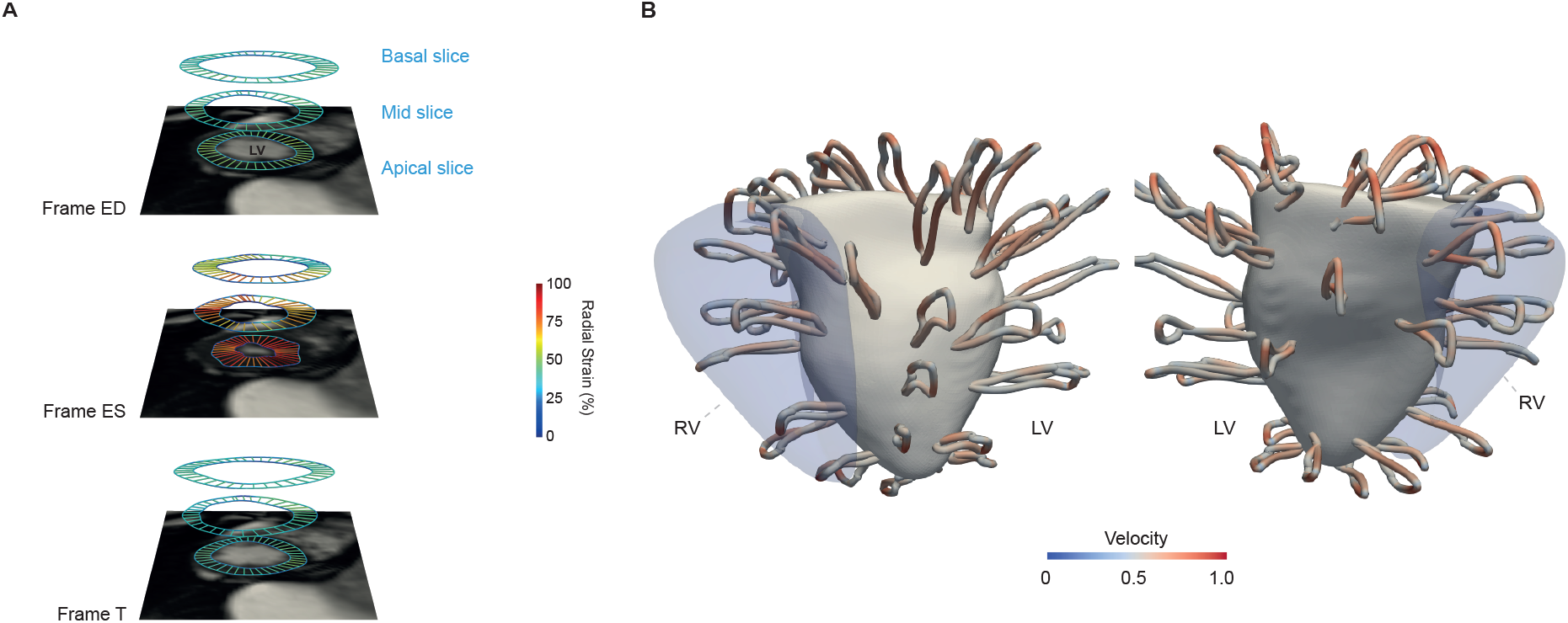
Segmentation and motion results. **A**, Example of deep learning left ventricular segmentation of short-axis cine images from the apical to the basal slices, across 50 temporal phases shown here at three time points. Here endocardial and epicardial surfaces are defined from which volumetric and functional parameters were calculated (value for regional radial strain shown). The motion fields and label maps were shape refined and registered to a cardiac atlas to produce a three dimensional mesh. **B**, Trajectory of left ventricular contraction and relaxation plotted as looped pathlines for a sub-sample of 100 points on a surface-shaded model of the heart at end systole. An outline of the right ventricle is shown for anatomic reference. The colour of the loops projecting from the surface represent the difference in velocity between healthy (n=5064) vs hypertensive (n=1330) participants. Individual-level dense motion coordinates were used as an input to the cardiac-age prediction networks. ED, end diastole; ES, end systole; LV, left ventricle; RV, right ventricle; T, temporal phase 1 - 50.

**Figure 4.**
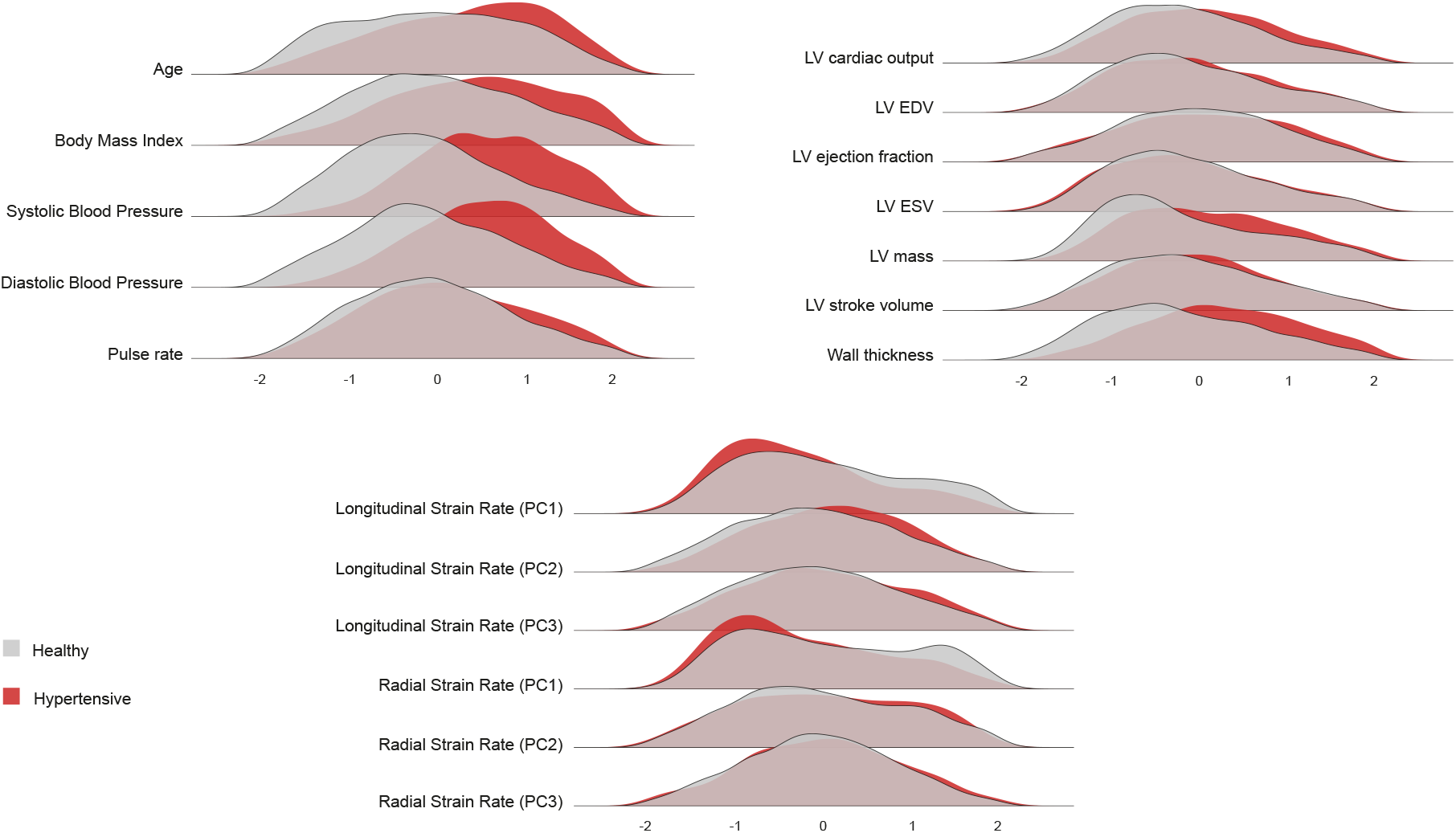
Population distribution of phenotypes. Ridge plots summarising the distribution densities for selected anthropometric, haemodymanic and cardiac measurements in healthy (n=5064) and hypertensive (n=1330) participants. LV, left ventricle; EDV, end diastolic volume; ESV, end systolic volume; PC, principal component.

### Cardiac motion graph representation

We modelled cardiac motion as a graph representation encoding spatio-temporal information. Each temporal phase is represented by an individual graph derived from the atlas-registered cardiac meshes consisting of a set of nodes with feature vectors capturing local geometry and positional information capturing temporal motion. The graph edges connecting nodes define spatial neighbourhood. The graphs were the input to a GNN trained to predict cardiac age. The GNN was a multi-graph network with convolutional submodels using GCNConv for graph convolutional layers.^9,10^ In Figure 5, we present an overview of the GNN architecture, with additional information in Supplementary Materials.

**Figure 5.**
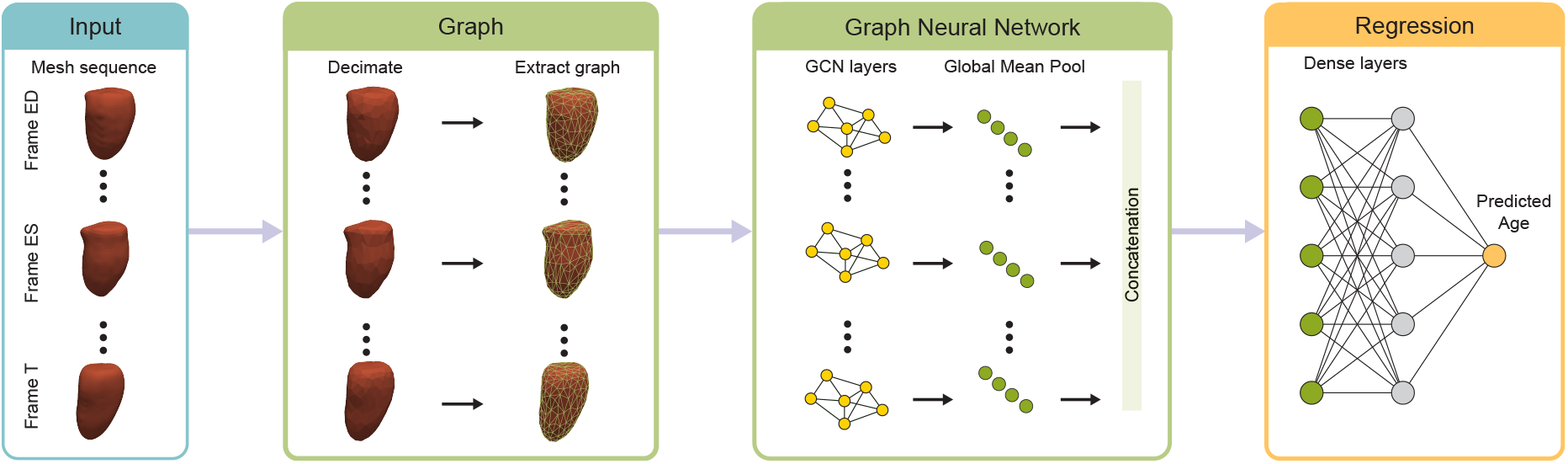
Overview of the prediction framework. High resolution time-resolved meshes of the left ventricle produced from the image analysis framework are first down-sampled using geometric decimation. Each frame is consumed by a graph convolutional submodel and transformed into a feature embedding using global average pooling. Cardiac age is then predicted from the concatenation of time-frame embeddings using a set of dense layers. (ED, end diastole; ES end systole; T, temporal phase 1 - 50; GCN, graph convolutional network.)

### Age prediction from cardiac motion

The performance comparison for age prediction using CatBoost, dense neural network and GNN models is shown in Table 3. All three models were trained on a normative cohort with healthy participants (n=4020), and evaluated on a holdout test set of healthy participants (n=1044). The CatBoost model comprised pre-defined imaging parameters and had the lowest performance for age prediction. Improved performance was achieved by a dense neural network which took time-resolved cardiac mesh coordinates as training data which were flattened in the input layer. Both models were outperformed by the GNN model, which was trained on graph representations capturing spatio-temporal relationships of cardiac motion. The GNN yielded a coefficient of determination (R^2^) of 0.35, a Pearson correlation coefficient (|r|) between predicted age and chronological age of 0.59, (*P* < 1 × 10^-16^) and a mean absolute error (MAE) of 4.74 years (Supplementary Materials).

**Table 3.**
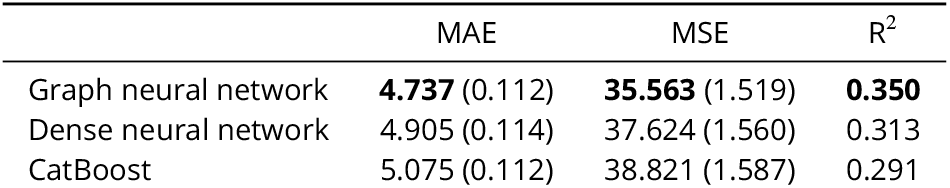
Age prediction performance comparison (trained on a set with n=4020 and evaluated in a holdout set with n=1044) across algorithms with standard errors presented in parentheses. MAE, mean absolute error; MSE, mean squared error; and R^2^, coefficient of determination.

### Effect of training data characteristics

We assessed a range of training sample sizes and degrees of mesh decimation (down-sampling) on the performance of the GNN (Table 4) in the same holdout set used for model comparison in Table 3. For these experiments, 100 hyperparameter searches were performed. A higher level of decimation decreases the performance (compared with the results in Table 3, where 90% decimation rate was used). Whilst reducing training set size significantly reduces predictive performance, a GNN model using only half of the dataset is sufficient to obtain a performance that outperforms CatBoost and dense neural network models using the whole dataset.

**Table 4.**
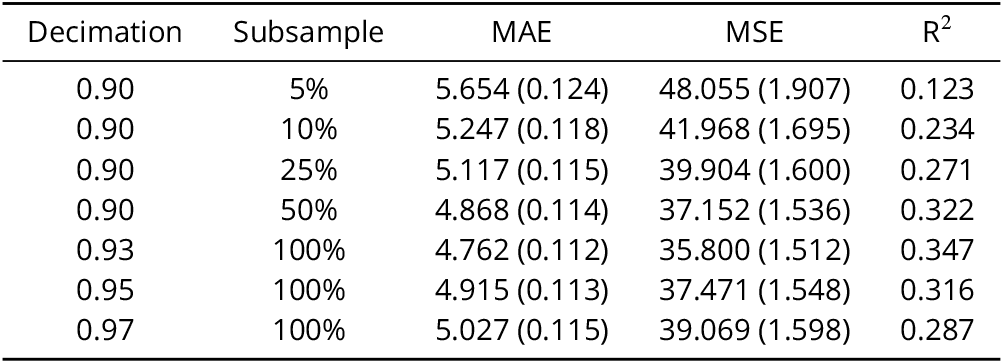
Age prediction performance comparison across GNNs with distinct levels of decimation and subsampling of the training set, i.e.: trained on a set of size 4020 multiplied by subsample factor (and rounded to nearest integer) and evaluated in a holdout set with n=1044. Standard errors are presented in parenthesis. MAE, mean absolute error; MSE, mean squared error; and R^2^, coefficient of determination. Smaller sample sizes and higher degrees of decimation both decrease the model performance.

| Decimation | Subsample | MAE | MSE | $R^2$ |
| --- | --- | --- | --- | --- |
| 0.90 | 5% | 5.654 (0.124) | 48.055 (1.907) | 0.123 |
| 0.90 | 10% | 5.247 (0.118) | 41.968 (1.695) | 0.234 |
| 0.90 | 25% | 5.117 (0.115) | 39.904 (1.600) | 0.271 |
| 0.90 | 50% | 4.868 (0.114) | 37.152 (1.536) | 0.322 |
| 0.93 | 100% | 4.762 (0.112) | 35.800 (1.512) | 0.347 |
| 0.95 | 100% | 4.915 (0.113) | 37.471 (1.548) | 0.316 |
| 0.97 | 100% | 5.027 (0.115) | 39.069 (1.598) | 0.287 |

### Age delta estimation

Age predictions for the healthy holdout set are shown in Figure 6A. Consistent with age prediction studies in other domains,^6,11^ a bias-correction step was performed to remove the effects of regression to the mean where age predictions are overestimated for younger participants and underestimated for older participants. Following bias-correction there was no residual correlation between cardiovascular age-delta and chronological age (|*r*| = 1.38 × 10^-8^, *P ≈* 1 for hypertensive participants, Figure 6B). Subsequently, we performed a 1:1 propensity matching procedure between healthy and hypertensive groups, matching on age and sex.

**Figure 6.**
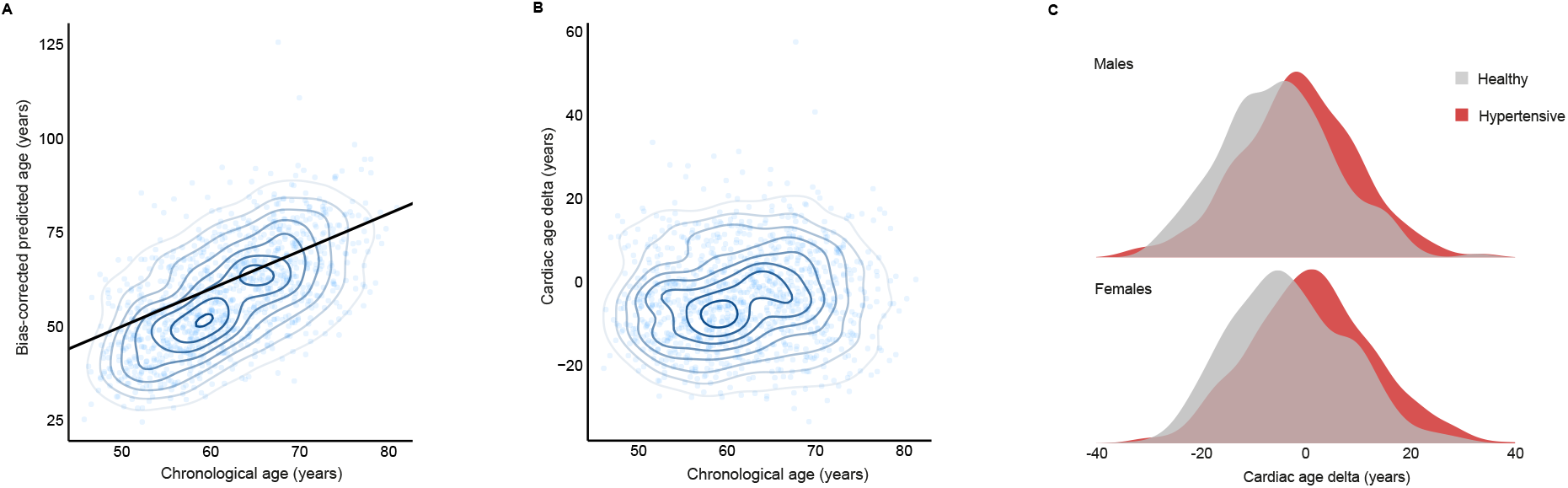
Predictions by the graph neural network. A, Predicted cardiac age and chronological age in healthy participants (n=1044). Ages jittered, linear model with 95% confidence intervals. B, Cardiac age-delta and chronological age, in healthy participants (n=1044). C, The gap between chronological age and predicted cardiac age (cardiac age-delta) is increased in subjects with hypertension. The grey shaded density plot represent age- and sex-matched controls with subjects with hypertension. The test samples comprised n=383 healthy males, n=612 hypertensive males; n=670 healthy females, n=718 hypertensive females.

Using linear regression to quantify the effect of hypertension on cardiac age-delta, we showed that hypertension was associated with an increase in predicted age in both males (+1.62 years, *P* < 1 × 10^-16^) and females (+4.19 years, *P* < 1 × 10^-16^) (Figure 6C and Supplementary Materials). We performed an additional linear regression including chronological age as an additional covariate, and found chronological age had a non-statistically significant coefficient (coefficient = 0.051, *P* = 0.176).

### Explainability of graph predictions

The ability to understand the predictions from the graph representations is valuable for physiological interpretation of the learned mapping from cardiac motion to ageing. Here, we employ the `GNNExplainer` algorithm to explain the predictions of the GNN model.^12^ Explanations are represented by node masks assigning an importance weighting to individual mesh nodes where the weights indicate a node’s influence for prediction.

To obtain a population-level explanation, we applied the `GNNExplainer` algorithm when processing an average temporal cardiac mesh sequence determined over all healthy participants. Extracted node masks at end diastole and end systole are shown in Figure 7. The node masks provide a compact visualization of age-related motion traits showing how these vary both anatomically and spatially in the heart. We also present alternative explainability visualizations based on saliency maps in Supplementary Materials.

**Figure 7.**
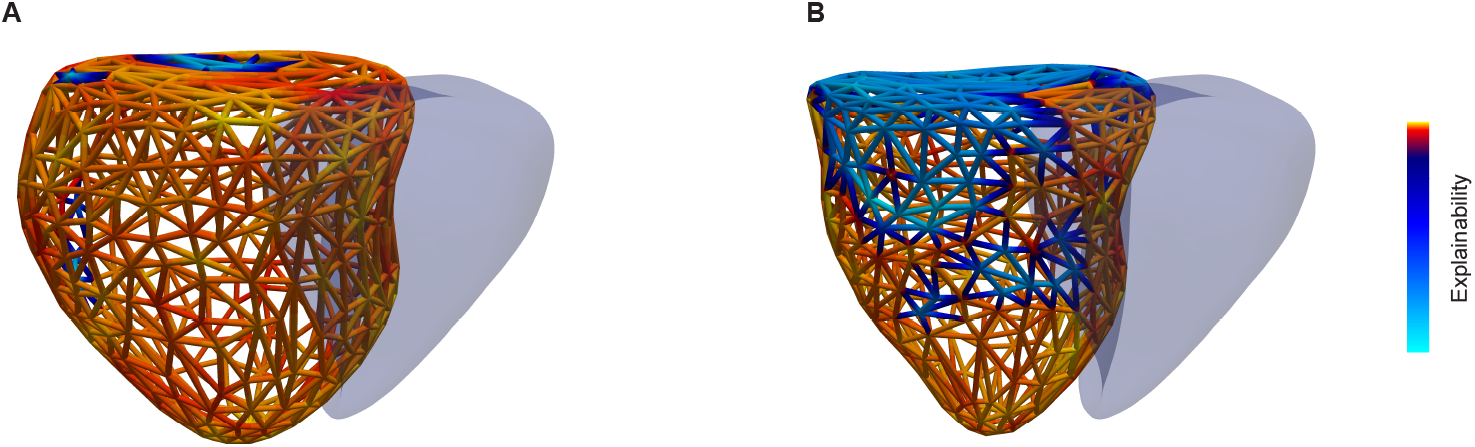
Explainability node masks. Visualization of the output of the `GNNExplainer` algorithm when processing an average temporal cardiac mesh sequence determined over all of healthy participants (n=5064) where **A** is at end diastole and **B** is at end systole. Red regions indicate higher node importance levels. An outline of the right ventricle is shown for anatomic reference. The models show how age prediction importance varies from base to apex across temporal frames.

## Discussion

Making inferences about future and current states of dynamic systems is a key challenge in biology and medicine. Here we show how the motion of the heart can be represented as a graph, where nodes are capturing spatio-temporal information with local geometric features and cardiac motion features encoded as relative 3D positions of mesh nodes over time. We apply a graph neural network model to the prediction task of a continuous outcome, learning multi-scale motion traits associated with biological ageing. This approach allows efficient prediction of a health state from high-dimensional meshes encoding the dynamic function of an organ system.

Cardiac ageing is a process that begins early in life and leads to a progressive change in structure and decline in function due to accumulated damage across diverse cell types, tissues and organs contributing to multi-morbidity. Cardiac ageing culminates in a final common pathway involving loss of tissue compliance which is manifest through diastolic dysfunction, tissue fibrosis and structural remodelling.^13,14^ While conventional image-derived phenotypes characterising these processes are associated with ageing,^6,15,16^ they rely on pre-defined parameters that may be weak estimators of the underlying biology and do not take advantage of the rich three dimensional structural and functional information encoded by cardiac imaging. Precision models of cardiac motion minimise information loss but pose challenges for how to efficiently predict an outcome from such high-dimensional inputs. Firstly, we ensured that the data were internally consistent, through shape refinement and atlas registration, such that cardiac motion is specified within a standard coordinate space with the same number and position of vertices in the heart.^3^ Previously reported data on mesh models encoding time-resolved motion have better predictive performance than low-dimensional features for semi-supervised outcome tasks in an auto-encoder framework but do not model spatial or temporal dependencies in the data.^4^ Graph neural networks address this by learning contextual spatial information, preserving the geometric and topological complexity of the latent structure. Graph neural networks have been used to quantify spatio-temporal patterns of cardiac motion from two dimensional anatomic contours,^17,18^ and here we represent the heart as a time-resolved three dimensional graph in a prediction task.

Visualisation of the three dimensional input data, using pathlines to project motion trajectories on to the surface of the heart, showed that there are complex differences in regional velocity profiles between healthy and hypertensive participants, while conventional volumetric and strain parameters can only capture global information from individual time points. This was reflected in the performance of CatBoost which only used conventional tabular data to predict cardiac age and had the lowest performance. The dense neural network, taking flattened mesh data as input, had improved predictive performance indicating the incremental information encoded by three dimensional motion. The GNN achieved the highest predictive performance by retaining spatio-temporal relationships of motion as a graph. These patterns could be visualised through explainabilty node masks and saliency maps indicating how regional motion contributes to the prediction. We also found that GNNs are efficient in terms of the scale of training data required to achieve the same performance as dense networks.

Cardiovascular disease is a leading cause of death, and premature ageing is a key risk factor for its development and progression. Predicting the difference between biological age and chronological age is an emerging tool for identifying the genetic basis of ageing and potential modifiable risk factors,^6^ but progress depends on efficient learning of age-related phenotypes. We show how raw imaging can be represented as a moving graph and used by a GNN for prediction in a fully automated, scalable and explainable framework. This approach has the potential to identify physiological mechanisms that define ageing and to screen individuals for any deviation from healthy ageing. In our exemplar, we show how hypertension accelerates ageing through complex bio-mechanical effects that are downstream of end-organ damage. More generally, we have demonstrated the power of graph models applied to dynamic biological systems for prediction tasks which could be generalised to problems where motion traits encode outcomes.

There are limitations to our study. Our GNN model was trained on the UKB population, which is predominantly European and has a higher participant uptake amongst females, those with higher socioeconomic standards and older individuals.^19^ The generalisability of our model could be extended by training it on a more diverse cohort that may exhibit different patterns of cardiac ageing. A more comprehensive model may also include multi-modal features,^20^ for instance electrocardiogram data, which are associated with ageing traits,^6^ and have been previously been used in classification tasks using GNNs.^21,22^In summary, this work demonstrates the predictive power of GNNs applied to graphs derived from routine medical imaging data. This represents a generalisable approach for modelling complex systems and specifically for assessing biological ageing. This approach could have broad applications in predicting outcomes and stratifying patients from three dimensional motion traits.

## Methods

### Datasets

The data used in this study were drawn from UKB which comprises approximately 500,000 community-dwelling participants aged 40–69 years who were recruited across the United Kingdom between 2006 and 2010.^7^ All participants provided written informed consent for participation in the study, which was approved by the National Research Ethics Service (11/NW/0382). Our study was conducted under terms of access approval number 40616. The UKB dataset was downloaded from https://biobank.ctsu.ox.ac.uk/. From 2014 a subset of participants was invited for multimodal imaging which included comprehensive CMR imaging performed to a standardised protocol.^8,23^ From 39,559 participants with CMR available we identified 5064 healthy participants. In order to define this population, we identified participants with a “good” or “excellent” self-reported level of health (UKB data-field 2178), and who were never-smokers (UKB data-field 22506), not taking regular medications (UKB data-field 20003), and did not have a diagnosis of cardiovascular, chronic respiratory and metabolic disease (e.g. hypercholesterolaemia, diabetes and obesity).^6^ This group, who would be expected to show a pattern of normal ageing, was used to train and evaluate our models.

In order to obtain a sample of participants with hypertension, disease definitions were treated as binary traits, and were defined within the UKB based on hospital records and self-reported data. We considered the participant to have hypertension if their records contained any of the ICD9 codes (4010, 4011, 4019, 4039), ICD10 codes (I100, I110, I119, I120, I129, I130, I131, I132, I139, I150, I51, I152, I159) or self-reported codes (1065, 1072, 1073) relevant to hypertension. Details of both healthy and hypertensive groups are shown in Table 1 and Figure 4.

The age of each participant is computed as the number of years between year of birth (UKB data-field 34) and date of CMR (UKB data-field 53). The sex of each participant is determined using UKB data-field 31.

### Image processing

#### Two dimensional segmentation

A standardised CMR protocol was followed to acquire two-dimensional (2D), retrospectively-gated cine imaging on a 1.5T magnet (Siemens Healthineers, Erlangen, Germany).^8^ Short-axis plane cine imaging comprised a contiguous stack of images from left ventricular base to apex, as well as long axis cine imaging in the two and four chamber views. Cine sequences consisted of 50 cardiac phases with a typical acquired temporal resolution of 31 ms. As previously described, these 2D cine images were segmented using a fully convolutional network (FCN), adapted from `VGG-16`,^24^ which learns image features from fine to coarse scales for predicting the label class at each pixel.^25^ Imaging phenotypes all underwent automated quality control prior to use in analysis.^26^ These label maps were used to derive biventricular and bi-atrial volumetric data with a performance comparable to human inter-observer variability.^25^ To assess dynamic function non-rigid image registration between successive frames enabled motion tracking on greyscale cine images.^27^ Circumferential (*E*_cc_) and radial (*E*_rr_) strains were calculated on the short axis cines by the change in length of respective line segments.^13^ Peak strain for each segment and global peak strain were then calculated. Strain rate was estimated as the first derivative of strain and peak early diastolic strain rate in radial (PDSR_rr_) and longitudinal (PDSR_ll_) directions were detected using local maxima.

#### Three dimensional models

Three dimensional (3D) representations of left ventricular shape and motion were derived from a shape-refined mesh fitted to the 2D segmentations and registered to an atlas template. Briefly, we use a multi-task deep learning network that simultaneously predicts segmentation labels and anatomical landmarks in CMR volumes.^28^ We introduce anatomical shape prior knowledge to the segmentation using atlas propagation from a cohort of high-resolution atlases. The pipeline outputs an accurate estimation of the true geometry of the ventricles from the 2D inputs that have low through plane resolution, incomplete coverage and inter-slice shifts (*Segmentation* in Figure 2). Subsequently, to process the segmentation results for motion tracking, we applied rigid, affine registration and non-rigid B-spline image registration on the obtained segmentation results for atlas alignment to enforce consistency of the mesh vertices (*Mesh fitting* in Figure 2). Then, motion tracking is achieved using non-rigid image registration between consecutive time frames using the `MIRTK` toolkit in forward and backward directions to avoid accumulation of registration errors. The displacement field is calculated by a weighted average of the forward field and backward field. For each cardiac phase we compute the displacement of each vertex from the previous and subsequent frame by assigning a weight as follows:

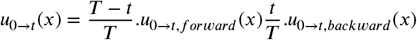

where *t*=1,…,50 shows the cardiac phase, and *T* is the maximum number of phases, i.e. 50 here. *u*_0*→t*_ is the displacement from frame 0 to frame *t* on pixel *x*. 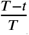 and 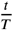 are the weights for the forward and backward displacement fields. Finally, we warp the reference frame (ED) with the final displacement field to obtain motion across all frames. As the motion field estimate lies within the reference space of each subject, we then warped the template mesh using the resulting rigid deformation and mapped it back to the template space (*Motion* in Figure 2). Fifty surface meshes, one for each temporal frame, were subsequently generated by applying the estimated motion fields to the warped template mesh accordingly (*Output* in Figure 2). Consequently, the surface mesh of each subject at each frame contained the same number of vertices (21510), which maintained their anatomical correspondence across temporal frames and participants.

### Graph neural network training and evaluation

Graph neural networks (GNNs) are machine learning algorithms specifically designed for the processing of graph-like data structures such as 3D meshes. Here, the cardiac meshes generated through our image pre-processing pipeline, consist of a set of nodes and edges between nodes naturally form a graph structure. Each cardiac time phase yields a graph, and each graph contains the same number of corresponding nodes as a result of the template mesh registration. The graph edges define the spatial neighborhood of nodes, and this information is used within the graph convolutional layers for message passing.^29^ The graph nodes can carry arbitrary feature vectors capturing relevant information for the prediction task at hand.

For our cardiac motion graph representation, a natural choice of features are the (x, y, z) mesh node coordinates. The relative change in node position captures temporal cardiac motion traits (Figure 3B). Additionally, we use geometric features to capture the local mesh geometry encoding both spatial (within mesh) and temporal (across meshes from time phases) anatomical information. Here, we employ the Fast Point Feature Histogram (FPFH) descriptors, originally proposed for mesh registration and recently shown to be powerful feature descriptors for shape classification.^10,30^ To determine the FPFH features on a mesh, first a point feature histogram must be generated. For each query point *p*_*r*_, all nearby points inside a 3D sphere with radius *r* and centering on point *p*_*r*_ are chosen (k-neighbor points); then, for every pair *p*_*r*_ and *p*_*k*_ in the k-neighbourhood points of *p*_*r*_ with their normals being estimated as *n*_*r*_ and *n*_*k*_. The point with the smaller angle between the line joining the pair of points and the estimated normals is chosen to be *p*_*r*_. Finally, a Darboux frame is defined as (*u* = *n*_*r*_, *v* = (*p*_*k*_ *− p*_*r*_) × *u, w* = *u* × *v*) and the angular variations of *n*_*r*_ and *n*_*k*_ are computed:

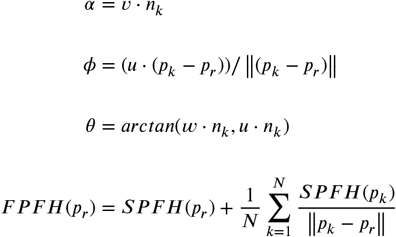

Where the *SP F H*(*p*_*r*_) (simple point feature histogram) is obtained by calculating the point features of each neighboring point *p*_*k*_.^31^ The FPFH feature vectors have length 34, which combined with the 3D positional information in terms of mesh node coordinates, yields node feature vectors of length 37. The input to our GNN model from a single subject is thus a set of 50 graphs with 37-dimensional node feature vectors.

We implemented our GNN model using the `PyTorch Geometric` Python package. The GNN architecture consisted of 50 submodels (one for each cardiac time phase). Each submodel consisted of two convolutional layers for which we employed the GCNConv operator.^9^ Submodels learned time phase specific feature embeddings using the two graph convolutions followed by a global average pooling across nodes. The resulting embeddings from each time phase were then concatenated and flattened into global feature representations which were processed by a prediction head with two dense layers. We used exponential linear units as activation functions after the first GCNConv layer in each submodel and the first dense layer of the prediction head. The GNN input data was preprocessed using the decimate method of `PyVista` Python package, with 90% decimation rate. In Figure 5, we present an overview of the GNN architecture, with additional details being presented in the Supplementary Methods.

We randomly split the healthy group into separate training (80%, n = 4020) and test (20%, n = 1044) sets. From the training set, a 10% validation holdout set (n = 403) was used for hyperparameter search, using the Python package `Optuna` with tree-structured Parzen estimator (TPE) search algorithm,^32^ where 300 models were trained. Additionally, from the remaining training set, a 10% additional validation holdout set was randomly taken to be used for early stopping evaluation of the GNN also using 300 for training.

In Table 5, we present details of the hyperparameter search space where the learning rate and weight decay hyperparameters are optimized in a log base 10 scale.

**Table 5.** Hyperparameter search space used for the graph neural networks. Learning rate and weight decay hyperparameters are optimized in a log base 10 scale. GCN, graph convolutional network.

|  | Lower bound | Upper bound |
| --- | --- | --- |
| Batch size | 10 | 20 |
| Weight decay | $10^{-15}$ | $10^{-5}$ |
| Learning rate | $10^{-5}$ | $10^1$ |
| First GCN layer output size | 10 | 50 |
| Second GCN layer output size | 10 | 50 |
| First dense layer output size | 100 | 1000 |

### Dense neural networks

As baseline for comparison, we trained dense neural networks, using the same decimated base meshes as used in the GNNs. An initial quadratic decimation was done on a template mesh, which as before, generates new points in space. Following this step, we use `MIRTK` registering and matching to find the nearest points in the original template set. We used the indices of those points to decimate all the participants’ meshes. Additional details of the architecture are in the Supplementary Methods.

The data setup for early stopping and hyperparameter search was identical to the GNN and, similarly, 300 models were trained and the model with the lowest mean squared error on the holdout dataset was chosen. In Table 6, we present details of the hyperparameter search space where the learning rate and weight decay hyperparameters are optimized in a log base 10 scale.

**Table 6.** Hyperparameter search space used for the dense neural networks. Learning rate and weight decay hyperparameters are optimized in a log base 10 scale.

|  | Lower bound | Upper bound |
| --- | --- | --- |
| Batch size | 50 | 110 |
| Weight decay | $10^{-10}$ | $10^{-1}$ |
| Learning rate | $10^{-5}$ | $10^1$ |
| First dense layer output size | 100 | 2000 |
| Second dense layer output size | 100 | 2000 |

### CatBoost

As another baseline for comparison, we trained a gradient boosting machine learning algorithm (CatBoost) on pre-defined tabular data. Gradient boosting algorithms are ensemble learning methods that successively train weak estimators (in the case of CatBoost, symmetrical decision trees) on the residuals of the weighted average of the weak estimators. The process of training extra weak estimators can continue until a fixed number of estimators are trained or until early stopping criteria are reached given that the loss on the training set should always decrease (or at least not increase), but the loss on the validation may eventually start increasing.

CatBoost was trained using 104 conventional CMR parameters as summarised in Table 1. These parameters are measures of left ventricular volume, function, mass and strain in longitudinal and radial directions. These low-dimensional features are implicit in the higher-dimensional time-resolved meshes used in our GNN model. CatBoost was trained using the default hyperparameter set (with automatic choice of learning rate and number of decision trees), except for early stopping rounds (patience) of 100, in this case, we used the reserved 10% hyperparameter search dataset as the evaluation for early stopping criteria.

### Cardiac age delta calculation

Tabular data have been used to compute “cardiovascular age-delta” in UKB using image-derived features of vascular and cardiac function.^6^ We follow a conceptually similar approach here using high-dimensional cardiac motion mesh data as inputs to the prediction network. We first predict age in the healthy group, and subtract this from the participants’ chronological age to obtain a cardiac age-delta, subsequently apply a bias-correction procedure,^11^ (Supplementary Materials) and then use this model to predict age in a group of participants with hypertension. Finally, we fit a linear regression with age-delta as the response variable and presence of disease as covariate. We confirmed there was no residual dependence of cardiac age delta on chronological age using linear regression.^33^

### Graph neural network interpretability

Saliency maps and the `GNNExplainer` algorithm were used to explore different explainability technqiues of our results.^12,34^ Saliency maps were originally proposed for image classification as a visualisation technique applied to one specific image and class at a time but can be adapted to regression and GNNs by leveraging the model’s ability to assign weights for each node of the input graph. We used the versatile implementation in the `Captum` Python package. `GNNExplainer`, on the other hand, was proposed to work directly on graph neural network models. The algorithm computes a node mask via iterative optimization identifying a node’s individual influence (or weight) on the final prediction. While saliency maps correspond to importance of specific graph edges, the `GNNExplainer`’s ability to highlight important nodes leads to better interpretability in our context where cardiac mesh nodes correspond to specific anatomical locations.

## Data Availability

All raw and derived data in this study are available from UK Biobank (http://www.ukbiobank.ac.uk/).

## Acknowledgments

The study was supported by the Medical Research Council (MC_UP_1605/13); British Heart Foundation (RG/19/6/34387, RE/18/4/34215), and the National Institute for Health Research (NIHR) Imperial College Biomedical Research Centre. This research has been conducted using the UK Biobank Resource under Application number 40616.

For the purpose of open access, the authors have applied a creative commons attribution (CC BY 4.0) licence to any author accepted manuscript version arising.

## Code Availability

The analysis code is freely available on GitHub (https://github.com/ImperialCollegeLondon/GNN_cardiac_ageing).

## Data Availability

All raw and derived data in this study are available from UK Biobank (http://www.ukbiobank.ac.uk/).

## Author contributions

M.H.de A.I. and M.S. performed the formal analyses and co-wrote the paper; W.B., M.J. and Q.M. performed the image analysis; N.S., A.G., B.G. performed additional data modelling. D.P.O’R conceived the study, managed the project and revised the manuscript. All authors reviewed the final manuscript.

