## Supplementary Materials for "Cardiac age prediction using graph neural networks"

### Participant characteristics

| Variables | Healthy (Mean) | Hypertension (Mean) | p-value | Adjusted p-value | Significance |
|---|---|---|---|---|---|
| Age at time of CMR (years) | 61.7 | 64.5 | 7.80x $10^{-36}$ | 3.90x $10^{-35}$ | **** |
| Race, Caucasian, n (%) | 0.9 | 0.9 | 0.027 | 0.03 | * |
| Body Mass Index (kg/m$^2$) | 24.3 | 25.5 | 1.80x $10^{-50}$ | 2.70x $10^{-49}$ | **** |
| Systolic Blood Pressure (mmHg) | 101.6 | 113 | 5.80x $10^{-09}$ | 1.20x $10^{-08}$ | **** |
| Diastolic Blood Pressure (mmHg) | 58.5 | 64 | 4.90x $10^{-07}$ | 7.30x $10^{-07}$ | **** |
| Pulse rate (bpm) | 69.5 | 71.8 | 4.10x $10^{-08}$ | 7.70x $10^{-08}$ | **** |
| LV end-diastolic volume (ml) | 143.8 | 146 | 0.028 | 0.03 | * |
| LV end-systolic volume (ml) | 58 | 58.4 | 0.51 | 0.51 | ns |
| LV stroke volume (ml) | 85.8 | 87.6 | 0.0017 | 0.0023 | ** |
| LV ejection fraction (%) | 60 | 60.4 | 0.016 | 0.02 | * |
| LV cardiac output (L/min) | 5.2 | 5.6 | 2.80x $10^{-19}$ | 8.50x $10^{-19}$ | **** |
| LV mass (g) | 79.5 | 86.8 | 3.00x $10^{-29}$ | 1.10x $10^{-28}$ | **** |
| Peak longitudinal strain rate (s$^{-1}$) | 1.8 | 1.5 | 3.80x $10^{-37}$ | 2.90x $10^{-36}$ | **** |
| Peak radial strain rate (s$^{-1}$) | -6.1 | -5.5 | 5.40x $10^{-15}$ | 1.30x $10^{-14}$ | **** |

**Table 1.** Comparison of attributes between healthy (n = 5064) and hypertensive groups (n = 1330), using unpaired T-tests and Benjamini-Hochberg correction. Abbreviations: CMR, Cardiac magnetic resonance; LV, left ventricle.

| Variables | Female (Mean) | Male (Mean) | p-value | Adjusted p-value | Significance |
|---|---|---|---|---|---|
| Age at time of CMR (years) | 61.3 | 62.4 | 1.20x $10^{-06}$ | 1.50x $10^{-06}$ | **** |
| Race, Caucasian (%) | 90 | 90 | 0.8 | 0.8 | ns |
| Body Mass Index (kg/m$^2$) | 23.9 | 24.9 | 1.30x $10^{-49}$ | 2.60x $10^{-49}$ | **** |
| Systolic Blood Pressure (mmHg) | 99.6 | 105 | 0.0014 | 0.0015 | ** |
| Diastolic Blood Pressure (mmHg) | 57.3 | 60.5 | 0.001 | 0.0012 | ** |
| Pulse rate (bpm) | 71.3 | 66.6 | 4.00x $10^{-33}$ | 7.10x $10^{-33}$ | **** |
| LV end-diastolic volume (ml) | 128.3 | 169.2 | 0 | 0 | **** |
| LV end-systolic volume (ml) | 50 | 71.2 | 0 | 0 | **** |
| LV stroke volume (ml) | 78.3 | 98 | 5.30x $10^{-288}$ | 1.90x $10^{-287}$ | **** |
| LV ejection fraction (%) | 61.1 | 58 | 5.20x $10^{-85}$ | 1.20x $10^{-84}$ | **** |
| LV cardiac output (L/min) | 4.9 | 5.8 | 8.30x $10^{-154}$ | 2.30x $10^{-153}$ | **** |
| LV mass (g) | 67.8 | 98.7 | 0 | 0 | **** |
| Peak longitudinal strain rate (s$^{-1}$) | 1.8 | 1.7 | 6.50x $10^{-20}$ | 9.00x $10^{-20}$ | **** |
| Peak radial strain rate (s$^{-1}$) | -6.3 | -5.7 | 3.70x $10^{-22}$ | 5.80x $10^{-22}$ | **** |

**Table 2.** Comparison of attributes between females (n = 3149) and males (n = 1915) in the healthy group, using unpaired T-tests and Benjamini-Hochberg correction. Abbreviations: CMR, Cardiac magnetic resonance; LV, left ventricle.

An overview of the participants used in this study are described in Table 1 and group comparisons are shown in supplementary tables 1, 2 and 3. 5064 “healthy” participants and 1330 participants diagnosed with hypertension were used in the study.

| Variables | Female (Mean) | Male (Mean) | p-value | Adjusted p-value | Significance |
|---|---|---|---|---|---|
| Age at time of CMR (years) | 64.6 | 64.4 | 0.69 | 0.81 | ns |
| Race, Caucasian (%) | 90 | 90 | 0.91 | 0.91 | ns |
| Body Mass Index (kg/m$^2$) | 25 | 26.1 | 8.20x $10^{-15}$ | 1.60x $10^{-14}$ | **** |
| Systolic Blood Pressure (mmHg) | 113.2 | 112.8 | 0.9 | 0.91 | ns |
| Diastolic Blood Pressure (mmHg) | 63.7 | 64.5 | 0.69 | 0.81 | ns |
| Pulse rate (bpm) | 73.8 | 69.4 | 3.60x $10^{-09}$ | 6.30x $10^{-09}$ | **** |
| LV end-diastolic volume (ml) | 128 | 167.1 | 2.80x $10^{-121}$ | 1.90x $10^{-120}$ | **** |
| LV end-systolic volume (ml) | 49.1 | 69.2 | 2.80x $10^{-99}$ | 1.30x $10^{-98}$ | **** |
| LV stroke volume (ml) | 78.8 | 97.9 | 6.60x $10^{-82}$ | 2.30x $10^{-81}$ | **** |
| LV ejection fraction (%) | 61.8 | 58.8 | 2.90x $10^{-20}$ | 6.70x $10^{-20}$ | **** |
| LV cardiac output (L/min) | 5.1 | 6.1 | 9.80x $10^{-51}$ | 2.80x $10^{-50}$ | **** |
| LV mass (g) | 73.1 | 102.8 | 9.20x $10^{-180}$ | 1.30x $10^{-178}$ | **** |
| Peak longitudinal strain rate ($s^{-1}$) | 1.6 | 1.5 | 0.00091 | 0.0013 | ** |
| Peak radial strain rate ($s^{-1}$) | -5.7 | -5.3 | 0.00023 | 0.00035 | *** |

**Table 3.** Comparison of attributes between females (n = 718) and males (n = 612) in the hypertensive group, using unpaired T-tests and Benjamini-Hochberg correction. Abbreviations: CMR, Cardiac magnetic resonance; LV, left ventricle.

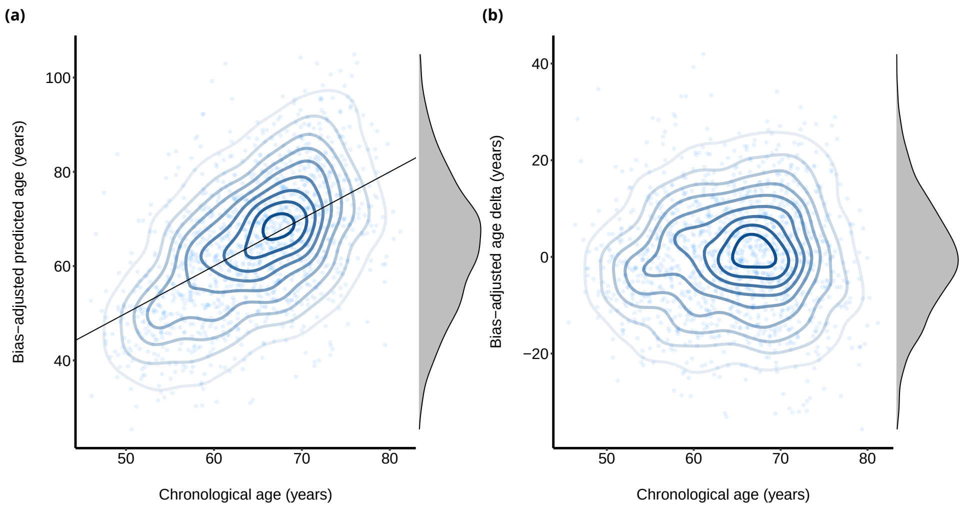


**Figure 1. The relationship between age delta, predicted age and chronological cardiovascular age in the hypertension cohort.** (a) Predicted and chronological cardiovascular age. (b) Delta and chronological cardiovascular age. Ages jittered, density contours, linear model with 95% confidence intervals, and a marginal density plot, (n=1330).

### Description of graph and dense neural networks

In Figure 2, we present the details of the graph neural network structure, where nodes are transformed using a concatenation of constant (preserve the original nodes as coordinates), position and Fast Point Feature Histograms features transformation, while edge attributes are generated as spherical coordinates of the edges.

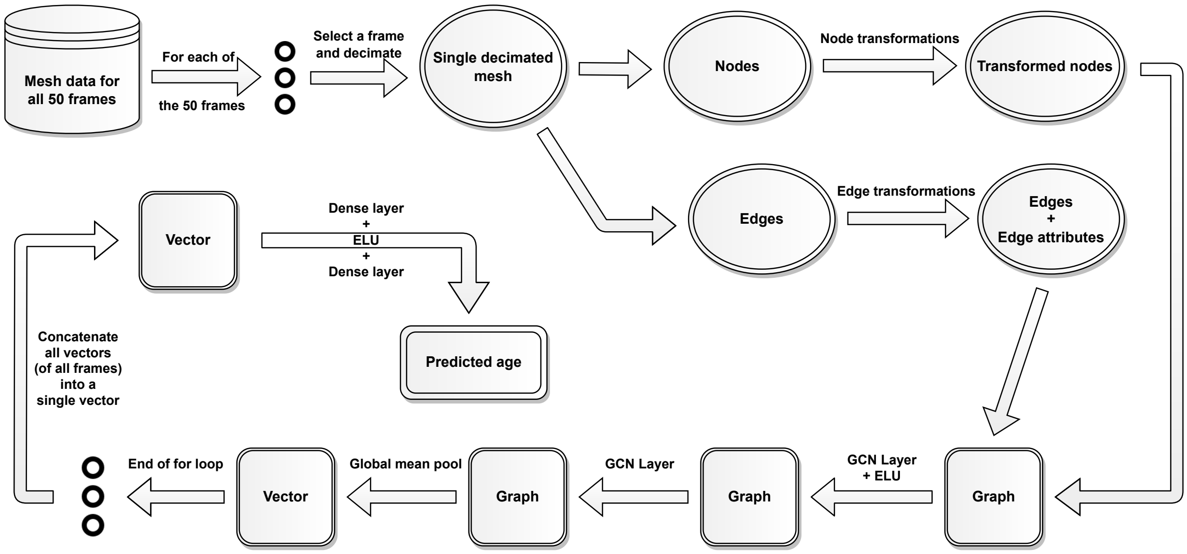


**Figure 2.** Details of the graph neural network structure, where nodes are transformed using a concatenation of constant, position and FPFH features transformations, while edge attributes are generated as spherical coordinates of the edges.

In Figure 3, we present the details of the dense neural network structure.

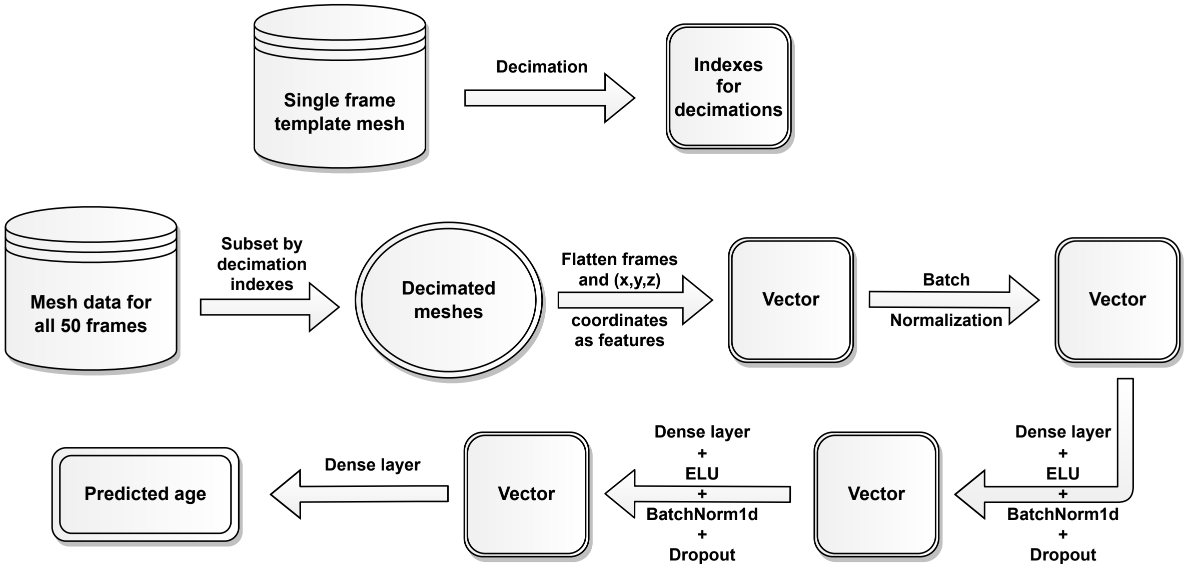


**Figure 3.** Details of the dense neural network structure.

### Age delta estimation

To obtain the age-delta estimate, we first predict age for both healthy and hypertension groups, and apply a bias correction procedure to the predictions (Algorithm 1). We next perform propensity matching by age and sex in the healthy and hypertension groups (n = 1044 in each group). We next fit a linear regression model where the predicted age is modelled as the response variable and the indicator function of whether the participant has hypertension or not is a covariate. For each covariate, we present the coefficient, standard error as well as the z-score, and the *P* value and confidence intervals. The heart of a person with hypertension has a biological age that is $\Delta$ years older than that of a healthy person, where $\Delta = 4.19$ for females and $\Delta = 4.19 - 2.57 = 1.62$ for males.

**Algorithm 1** Bias correction procedure

```
input ground_truth, prediction
output corrected_prediction
procedure Bias correct
    reg = LinearRegression()
    reg.fit(x=ground_truth, y=prediction)
    intercept = reg.intercept
    coefficient = reg.coefficient
    corrected_prediction = (prediction - intercept) / coefficient
end procedure
```

**Table 4. Age delta regression results.** Summary statistics for the ordinary least squares linear regression performed to assess the effect of hypertension on cardiac age-delta. The response variable is the cardiac age-delta. Coefficient, standard errors as well as the z-score, and the *P* value and 95% confidence interval limits displayed. n = 2088.

| | Coefficient | Standard Error | z-score | P > \|z-score\| | Lower confidence interval | Upper confidence interval |
|---|---|---|---|---|---|---|
| **Intercept** | -2.826 | 0.397 | -7.110 | $< 10^{-16}$ | -3.604 | -2.047 |
| **Hypertension** | 4.191 | 0.506 | 8.284 | $< 10^{-16}$ | 3.200 | 5.183 |
| **Sex** | -2.570 | 0.509 | -5.053 | $< 10^{-16}$ | -3.566 | -1.573 |

**R-squared:** 0.038 **Adj. R-squared:** 0.037

### Saliency maps

In Figure 4, we present a visualisation of the explainability algorithm using saliency maps when predicting age for an average mesh determined over all of healthy participants versus an average mesh determined over all of hypertensive participants. Darker regions indicate higher importance levels.

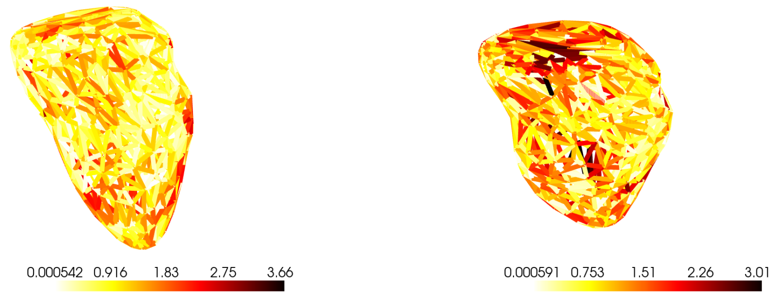


(a) (b)

**Figure 4. Explainability using saliency maps.** Explanability algorithm using saliency maps when predicting age for averages meshes over **A** all of healthy participants and **B** all of hypertensive participants. Darker regions indicate higher importance levels.
